# Eight-year Combined Diet and Physical Activity Intervention Affects Serum Metabolites during Childhood and Adolescence: A Nonrandomized Controlled Trial

**DOI:** 10.1101/2024.04.01.24305105

**Authors:** Iman Zarei, Aino-Maija Eloranta, Anton Klåvus, Juuso Väistö, Marko Lehtonen, Santtu Mikkonen, Ville M. Koistinen, Taisa Sallinen, Eero A. Haapala, Niina Lintu, Sonja Soininen, Mustafa Atalay, Ursula Schwab, Seppo Auriola, Marjukka Kolehmainen, Kati Hanhineva, Timo A. Lakka

## Abstract

**Background:** Molecular mechanisms underlying the beneficial effects of long-term lifestyle interventions on cardiometabolic health during childhood and adolescence remain largely unknown. Such information would provide valuable insights into the prevention of cardiometabolic diseases since childhood. We therefore studied for the first time the effects of a long-term diet and physical activity (PA) intervention on serum metabolites in a general population of children.

**Methods:** We carried out an 8-year, nonrandomized, controlled trial in a population sample of 490 prepubertal children (257 girls, 233 boys) aged 6–9 years followed up until adolescence. We allocated the children to a combined diet and PA intervention group and a control group. We performed a non-targeted liquid chromatography–mass spectrometry (LC-MS) metabolomics analysis of fasting serum samples at baseline, two years, and eight years. We analyzed the intervention effects on serum metabolites using linear mixed-effects models adjusting for sex and age.

**Results:** The intervention had effects on 80 serum metabolites over two years, 17 of these metabolites being affected by the interevention until eight years. The intervention had effects on several fatty amides (such as palmitic amide, linoleamide, oleamide, elaidamide, capsiamide, myristamide, palmitoleamide, docosanamide, and erucamide), unsaturated fatty acids (such as 12-hydroxyheptadecatrienoic acid, hydroxyeicosatetraenoic acid, hydroxyoxohexadecanoic acid, and oxotetradecenoic acid), and acylcarnitines (such as octanoyl-L-carnitine, decatrienoylcarnitine, and valerylcarnitine) as well as many phospholipids and sterols over two years. Moreover, the intervention affected several gut-microbiota-derived metabolites (such as hydroxyferulic acid, hippuric acid, indolepropionic acid, pyrocatechol sulfate, 3-carboxy-4-methyl-5-pentyl-2-furanpropanoic acid, *p*-cresol sulfate, indolelactic acid, and 3,4-dimethyl-5-pentyl-2-furanpropanoic acid), amino acids (such as methoxybenzenepropanoic acid, glutamic acid, taurine, and hydroxyisoleucine), and purine metabolites (such as guanosine, inosine, xanthine, and hypoxanthine) over two years.

**Conclusions:** The diet and PA intervention had long-term effects on numerous serum metabolites that could influence cardiometabolic health since childhood. The intervention effects were most pronounced on serum fatty amides, but the intervention also affected other potentially important serum lipids, including fatty acids, acylcarnitines, phospholipids, and sterols, as well as serum gut-microbiota-derived metabolites, amino acids, and purine metabolites. These metabolites could be molecular mechanisms underlying the beneficial effects of long-term lifestyle interventions on cardiometabolic health since childhood.

**Trial Registration:** ClinicalTrials.gov NCT01803776. Registered 01 October 2007, https://clinicaltrials.gov/study/NCT01803776

## INTRODUCTION

The alarmingly high and rapidly increasing prevalence of childhood overweight and associated cardiometabolic risk factors is an important clinical, public health, and societal problem worldwide [1–3]. A major concern is that overweight and other cardiometabolic risk factors worsen since childhood and markedly increase the risk of cardiometabolic diseases in adulthood [4–8]. Pathophysiological processes underlying the development of cardiometabolic diseases begin in childhood or even during the fetal period [1, 2, 9] which emphasizes the need for preventing these diseases since childhood [1, 9–12].

Long-term diet and physical activity (PA) interventions have been found to decrease adiposity [13, 14], reduce insulin resistance [15, 16], and improve blood lipids [17, 18] in general populations of children. However, molecular mechanisms underlying the beneficial effects of lifestyle interventions on cardiometabolic health during childhood and adolescence remain largely unknown. A nontargeted liquid chromatography - mass spectrometry (LC-MS) metabolomics analysis of blood samples is a sensitive, high-throughput method to detect endogenous metabolites and exogenous compounds [19] that can be used to reveal molecular mechanisms of the beneficial effects of lifestyle interventions on cardiometabolic health in all age groups. Diet and PA interventions using the nontargeted LC-MS metabolomics analysis have been shown to affect numerous blood metabolites among adults [20–22]. Part of these metabolites, such as indolepropionic acid and hippuric acid, have been proposed as molecular mechanisms underlying the beneficial effects of lifestyle interventions on glucose metabolism and the risk of developing type 2 diabetes [20–22].

There are no reports on the effects of long-term lifestyle interventions on blood metabolites using the nontargeted LC-MS metabolomics analysis in general populations of children followed up until adolescence. Such scientific evidence would increase our understanding of molecular mechanisms underlying the beneficial effects of lifestyle interventions on cardiometabolic health during childhood and adolescence and would thereby provide valuable insights into the prevention of cardiometabolic diseases since childhood. We therefore studied for the first time the effects of a long-term diet and PA intervention on a wide spectrum of serum metabolites using the nontargeted LC-MS metabolomics analysis [19] in a population sample of children followed up for eight years until adolescence.

## METHODS

### Study Design and Participants

The Physical Activity and Nutrition in Children (PANIC) study is an 8-year, nonrandomized, controlled trial to investigate the long-term effects of a combined diet and PA intervention on cardiometabolic risk factors in a general population of children from the city of Kuopio, Finland (<u>NCT01803776</u>) [16–18, 23, 24]. The Research Ethics Committee of the Hospital District of Northern Savo approved the study protocol in 2006 and 2015 (Statements 69/2006 and 422/2015). The caregivers gave their written informed consent, and the children provided their assent to participation. Moreover, the caregivers and adolescents gave their written informed consent before the 8-year examinations. The PANIC study has been carried out in accordance with the principles of the Declaration of Helsinki as revised in 2008.

We have described the study design, the study setting, the recruitment of participants, the inclusion and exclusion criteria, the reasons for dropping out, and the assessments in detail previously [16–18, 23] and also provide this information in **Figure 1**. In brief, we invited 736 children 6–9 years of age who started the first grade in 16 primary schools of the city of Kuopio in 2007–2009 to participate in the study. Altogether, 512 children (248 girls, 264 boys) who accounted for 70% of those invited, accepted the invitation and attended the baseline examinations between October 2007 and December 2009. We excluded eight children at baseline; six children were either owing to their physical disabilities that could hamper participation in the intervention or withdrawal of the families because they had no time or motivation to attend the study, and two children whose parents or caregivers later withdrew their permission to use these data in the study. The final study sample thus included 504 children at baseline.

**Figure 1.**
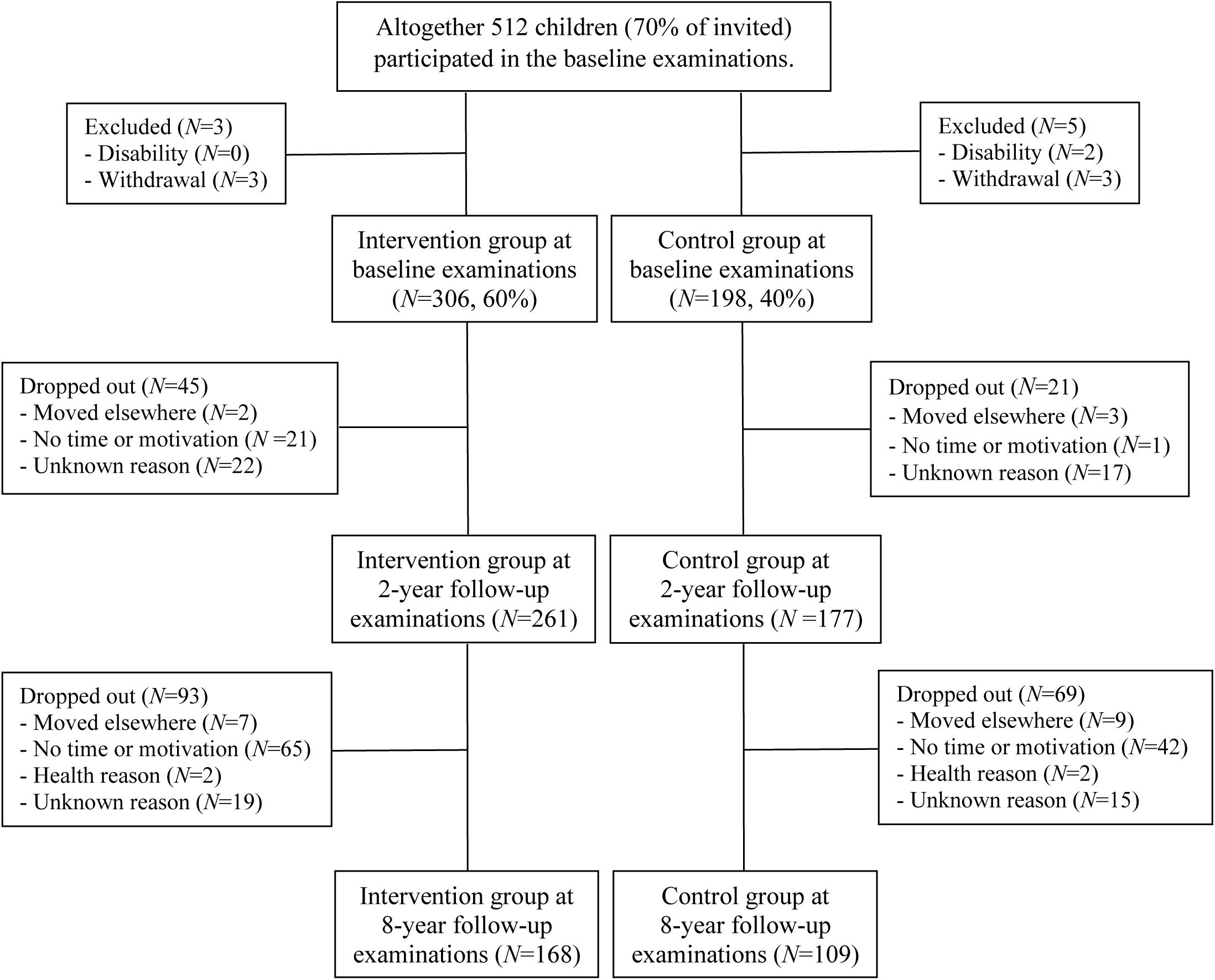
Flowchart of the Physical Activity and Nutrition in Children (PANIC) study

We allocated the 504 children from nine schools to a combined diet and physical activity intervention group (306 children, 60%) and the children from seven schools to a control group (198 children, 40%) (**Figure 1**) to avoid contamination in the control group by any local or national health promotion programs that could have been initiated in the study region during the study. We also proportionally matched the intervention and control group according to the location of the schools (urban vs rural) to minimize sociodemographic differences between the groups. We included more children in the intervention group than in the control group because of a larger number of dropouts expected in the intervention group and to retain a sufficient statistical power for comparison between the groups. The children, their parents or caregivers, or people carrying out the examination visits or doing the measurements were not blinded to the group assignment.

Of all 504 children who participated in the baseline examinations, 438 (87%) attended the 2-year examinations, and 277 (55%) attended the 8-year examinations (**Figure 1**). At the 2-year examinations, 261 children were from the intervention group (85%) and 177 children were from the control group (89%). At the 8-year examinations, 169 (55%) adolescents were from the intervention group and 108 (55%) adolescents were from the control group. Those who participated in the 2-year or 8-year examinations did not differ in age, height-SDS, BMI-SDS, or the distribution of sex or study groups at baseline from those who dropped out.

We excluded 14 children who had entered clinical puberty by the baseline examinations from the present statistical analyses to minimize possible confounding by metabolic changes caused by puberty. After excluding these children, the number of participants in these statistical analyses was 490 (257 girls, 233 boys) at the baseline examinations, 424 (219 girls, 205 boys) at the 2-year examinations, and 270 (122 girls, 148 boys) at the 8-year examinations.

### Diet and Physical Activity Intervention

The goals of the individualized and family-based diet and physical activity intervention were to 1) decrease the consumption of significant sources of saturated fat and particularly high-fat dairy and meat products, 2) increase the consumption of significant sources of unsaturated fat and particularly high-fat vegetable oil-based margarines, vegetable oils, and fish, 3) increase the consumption of vegetables, fruits, and berries, 4) increase the consumption of significant sources of fiber and particularly whole grain products, 5) decrease the consumption of significant sources of sugar and particularly sugar-sweetened beverages, sugar-sweetened dairy products, and candy, 6) decrease the consumption of significant sources of salt and the use of salt in cooking, 7) increase total physical activity by emphasizing its diversity, 8) decrease total and particularly screen-based sedentary behavior, and 9) avoid excessive energy intake. The goals of the diet and physical activity intervention were based on the national nutrition and physical activity recommendations.

We have described the individualized and family-based diet and PA intervention in detail earlier [16–18, 23, 24]. In brief, the intervention during the first two years consisted of six intervention visits that occurred 0.5, 1.5, 3, 6, 12, and 18 months after the baseline examinations [16, 17, 24]. Each intervention visit included 30–45 minutes of dietary counselling and 30–45 minutes of physical activity counselling for the children and their parents or caregivers. The children and their parents or caregivers received individualized advice from a clinical nutritionist and a specialist in exercise medicine on how to improve the diet, increase physical activity, and decrease sedentary time of the children in everyday conditions. Each visit had a specific topic on diet, physical activity, and sedentary behavior according to the intervention goals and included practical tasks on these topics for the children. The children and their parents or caregivers also received fact sheets on diet, physical activity, and sedentary time, as well as verbal and written information on opportunities for exercising in the city of Kuopio. Some material support was also given for physical activity, such as exercise equipment and allowance for playing indoor sports. Of the 306 children in the intervention group who attended the baseline examination, 266 (87%) participated in all six visits, 281 (92%) in at least five visits, and 295 (96%) in at least four visits. The children in the intervention group, particularly those who did not attend organized sports or exercise, were also encouraged to participate in after-school exercise clubs organized at the nine schools by trained exercise instructors of the PANIC study. The children in the control group were not allowed to attend these exercise clubs to avoid a non-intentional intervention in the control group.

After the 2-year follow-up examinations, the diet and physical activity intervention was mild and was continued until the 8-year examinations [23]. The intervention included individualized diet and physical activity counseling sessions 3, 5, 6, and 7 years after baseline and a group-based counseling session at schools 4 years after baseline. The participants were able to attend the counseling sessions occurring 3, 5, 6, and 7 years after baseline with or without their parents, who were given individualized counseling in a separate room. Each of these counseling sessions had a specific topic on diet, physical activity, and sedentary behavior according to the intervention goals. The group-based counseling session at schools 4 years after baseline included an active lesson with practical tasks.

The children and their parents in the control group received general verbal and written advice on health-improving diet and physical activity at baseline, but they were not given active intervention.

### Assessment of Diet and Physical Activity

We have described the assessment of dietary factors and physical activity in detail earlier [16, 17, 23, 24]. In brief, the consumption of food and drinks were assessed using 4-day food records. The caregivers recorded all food and drinks consumed by their children at baseline and 2-year examinations, whereas the adolescents recorded their food and drink consumption at the 8-year examinations. We calculated food consumption and nutrient intake using the Micro Nutrica^®^ dietary analysis software, Version 2.5, which is based on detailed information about the nutrient content of foods in Finland and other countries [25, 26]. Moreover, the clinical nutritionists updated the software by adding new food items and products with their actual nutrient content based on new data in the Finnish food composition database (https://fineli.fi/fineli/fi/index) or received from the producers. We assessed total physical activity energy expenditure, light, moderate, and vigorous physical activity, as well as total sedentary time using individually calibrated combined heart rate and body movement monitoring [16]. Average total physical activity energy expenditure was calculated in kJ × kg^-1^ daily. Light, moderate, and vigorous physical activity were defined as time spent at intensity >1.5 and ≤4.0 metabolic equivalents (METs), >4.0 and ≤7.0 METs, and >7.0 METs, respectively, by defining one MET as an energy expenditure of 71 J × kg^-1^ × min^-1^ or oxygen uptake of 3.5 ml × kg^-1^ × min^-1^. Total sedentary time was defined as time spent at intensity ≤1.5 METs excluding sleep.

### Assessment of Body Composition and Puberty

We assessed body height using a wall-mounted stadiometer and body weight using the InBody^®^ 720 bioelectrical impedance device (Biospace, Seoul, South Korea), with the weight assessment integrated into the system, in the morning after the participants had fasted for 12 hours [16]. We computed age- and sex-standardized BMI-SDS using Finnish references [27].

We defined overweight and obesity using the International Obesity Task Force criteria, corresponding to an adult BMI cut-point of 25 for overweight and 30 for obesity [28]. Research physicians carried out a medical examination, assessed pubertal status, and defined clinical puberty using stages described by Tanner as breast development stage ≥ 2 for girls and testicular volume ≥ 4 mL assessed using an orchidometer for boys [29, 30].

### Sample Preparation for Metabolomics Analyses

We have described the metabolomics analysis and the identification of serum metabolites in detail earlier [19]. Briefly, we performed the nontargeted LC-MS metabolomics analysis of fasting serum samples at the Metabolomics Center, Biocenter Kuopio, Finland. Fasting serum samples were randomized before the LC-MS analysis. After the samples were thawed entirely in ice bath for three hours, 100 µl of each sample was mixed with 400 µl of acetonitrile and centrifuged (2500 rpm, 4 °C, 5 minutes) on a 96-well filter plate (Captiva ND Plate 0.2 µm PP, Agilent Technologies, USA) to precipitate proteins from the sample [19]. The samples were pipette mixed with four pipette strokes to thoroughly precipitate serum proteins. The samples were centrifuged (700 × g for 5 min, 4° C) and the supernatants were collected to a 96-well plate (96 DeepWell PP Plate, Thermo Fisher Scientific Nunc, Rochester, NY, USA) which was covered (96 Well Cap Natural, Thermo Fisher Scientific Nunc A/S, Roskilde, Denmark). A small part, approximately 2 µl, of the 187 randomly chosen protein precipitated study samples, was pooled and used as a quality control (QC) sample. These QC samples were injected at the beginning of the sequence to equilibrate the analytical platform and then after every 12^th^ sample throughout the analysis. In addition, a solvent blank was prepared and injected at the beginning of the sequence.

### Liquid Chromatography - Mass Spectrometry Analyses

The LC-MS analysis for the non-targeted metabolite profiling was carried out at the LC-MS metabolomics center (Biocenter Kuopio, University of Eastern Finland, Finland). To meet the wide diversity of sample components, all samples were analyzed using two different chromatographic techniques, i.e., reversed phase and hydrophilic interaction chromatography (HILIC) for amphiphilic and hydrophilic metabolites, respectively. In addition, the data were acquired in both electrospray ionization (ESI) polarities, i.e., ESI positive (ESI+) and ESI negative (ESI-).

#### Liquid Chromatography - Mass Spectrometry Analyses of Amphiphilic Metabolites

The analysis of amphiphilic metabolites was carried out using an ultra-high performance liquid chromatography (Vanquish Flex UHPLC system, Thermo Scientific, Bremen, Germany) coupled online to a high-resolution mass spectrometry (HRMS, Q Exactive Classic, Thermo Scientific, Bremen, Germany). Samples were analyzed using a reversed phase technique. The sample solution (2 µl) was injected onto a RP column (Zorbax Eclipse XDBC18, 2.1 × 100mm, 1.8 μm, Agilent Technologies, Palo Alto, CA, USA) that was kept at 40 °C. The mobile phase, delivered at 400 μL/min, consisted of water (eluent A) and methanol (eluent B), both containing 0.1 % (v/v) of formic acid. The following gradient profile was used: 0–10 min: 2 to 100% B, 10–14.50 min: 100% B, 14.50–14.51 min: 100 to 2% B; 14.51–20 min: 2% B. The sample tray was at 10 °C during these analyses. Mass spectrometry was equipped with a heated ESI. The positive and negative ionization modes were used to acquire the data in centroid mode. The following ESI source settings were utilized; spray voltage 3.0 kV for positive and negative ionization modes, sheath gas (20), auxiliary gas (5), and sweep gas (1) (flow rates as arbitrary units for ion source). The capillary temperature and the probe heater temperature were set to 350 °C and 400 °C, respectively. The S-lens RF level was set to 50 V. A full scan range from 120 to 1,200 (m/z) was used with the resolution of 70,000 (m/Δm, full width at half maximum at 200 u). The injection time was set to 200 ms, and Automated Gain Control (AGC) was targeted at 1,000,000 ions.

For product ion scan (MS/MS) experiments, the Q-Exactive spectrometer was used with the same source parameters and chromatography conditions as described above. Two scan events were used: (a) an MS scan with a mass resolution power, AGC target and maximum injection time set to 70,000 (m/Δm, full width at half maximum at 200 u), 1,000,000 ions and 100 ms, respectively, and (b) an MS/MS scan (in HCD mode) at a normalized collision energy ranging from 20 to 40 %, depending on the molecule, with a mass resolution power, AGC target, maximum injection time, isolation window set to 17,500 (m/Δm, full width at half maximum at 200 u), 50,000, 50 ms and 1.5 m/z, respectively. Loop count was 3, apex trigger 0.2 to 3 s, and dynamic exclusion 15 s. The data for MS/MS experiments was collected in profile mode. The detector was calibrated before the sample sequence and subsequently operated at high mass accuracy (<2 ppm). Continuous mass axis calibration was performed by monitoring reference ions m/z 214.08963 in the positive ionization mode.

#### Liquid Chromatography - Mass Spectrometry Analyses of Hydrophilic Metabolites

For the analysis of hydrophilic compounds, an ultra-high performance liquid chromatography (1290 LC system, Agilent Technologies, Waldbronn, Karlsruhe, Germany) coupled online to a high-resolution mass spectrometry (HRMS, 6540 UHD accurate-mass quadrupole-time-of-flight (qTOF) mass spectrometry, Agilent Technologies, Waldbronn, Karlsruhe, Germany) was used. Samples were analyzed using a hydrophilic interaction chromatography technique (HILIC). The sample solution (2 µl) was injected onto a column (HILIC, Acquity UPLC^®^ BEH Amide 1.7 µm, 2.1×100 mm, Waters Corporation, Milford, MA, USA) that was kept at 45 °C. Mobile phases, delivered at 600 μl/min, consisted of 50% (v/v) (Eluent A) and 90% (v/v) (Eluent B) acetonitrile, respectively, both containing 20 mM ammonium formate (pH 3). Following gradient profile was used: 0–2.5 min: 100% B, 2.5–10 min: 100% B → 0% B, 10–10.1 min: 0% B → 100% B; 10.1–12.5 min: 100% B. The total runtime was 12.5 min. The sample tray was at 10 °C during these analyses.

Mass spectrometry was equipped with a heated ESI source, operated in both positive and negative ionization modes. The source used the following conditions: drying gas temperature 325 °C and a flow of 10 l/min, sheath gas temperature 350 °C and a flow of 11 l/min, nebulizer pressure 45 psi, capillary voltage 3500 V, nozzle voltage 1000 V, fragmentor voltage 100 V, and skimmer 45 V. Nitrogen was used as the instrument gas. For data acquisition, a 2 GHz extended dynamic range mode was used in both positive and negative ion modes from 50 to 1600 (m/z). Data was collected in the centroid mode at an acquisition rate of 2.5 spectra/s (i.e., 400 ms/spectrum) with an abundance threshold of 150.

For automatic data dependent MS/MS analyses, the precursor isolation width was 1.3 Da, and from every precursor a scan cycle of 4 most abundant ions were selected for fragmentation. These ions were excluded after 2 product ion spectra and released again for fragmentation after a 0.25 min hold. Precursor scan time was based on ion intensity, ending at 20,000 counts or after 300 ms. Product ion scan time was 300 ms. Collision energies were 10, 20, and 40 V in subsequent runs.

The TOF was calibrated before the sample sequence and subsequently operated at high accuracy (<2 ppm). Continuous mass axis calibration was performed by monitoring two reference ions from an infusion solution throughout the runs. The reference ions were m/z 121.050873 and m/z 922.009798 in the positive mode and m/z 112.985587 and m/z 966.000725 in the negative mode. Both high resolution mass spectrometries were equipped with a heated electrospray source, and the data was acquired in positive and negative ionization modes. Data-dependent MS/MS data were acquired at the beginning and end of the analysis from the quality control samples.

### Metabolomics Data Analyses

Data processing was performed separately for each of the four analytical modes. The molecular features were obtained using the MS-DIAL software [31]. The data were preprocessed using the R software (R Core Team, 2023, https://www.R-project.org), where the molecular features were corrected for the drift pattern caused by the LC-MS procedures, and feature quality was assessed based on the quality control samples [19]. First, the features were log-transformed. Regularized cubic spline regression was fit separately for each feature on the quality control samples. The smoothing parameter was chosen from an interval between 0.5 and 1.5 using leave-one-out cross validation to prevent overfitting. Features were kept if their non-parametric relative standard deviation and their D-ratio were below 20% and 40%, respectively. In addition, all the features with a relative standard deviation, a non-parametric relative standard deviation, and basic D-ratio below 10% were kept. This additional condition prevents the flagging of features with very low values in all but a few samples. These features tend to have a very high value of non-parametric D-ratio, since the median absolute deviation of the biological samples is not affected by the large concentration in a handful of samples, causing the non-parametric D-ratio to overestimate the significance of random errors in measurements of quality control samples. Thus, other quality metrics were applied with a conservative limit of 0.1 to ensure that only good quality features were kept this way [19, 32]. The Metabolomics data were missing at random. Missing values were imputed using random forest imputation in two phases. First, only the good quality features were imputed to prevent the flagged features from affecting the imputation. Thereafter, the flagged features were imputed [33]. After imputation, all batches of a single mode were combined, and the median of each quality metric across batches was chosen as an overall quality metric. The preprocessed data was entered in statistical analyses, and molecular features that were statistically significantly affected by the intervention were addressed for metabolite identification following the 4-level metabolite annotation scheme proposed by the Metabolomics Standards Initiative [34].

### Compound Identification

The annotation of compounds based on spectral database searches was performed in MS-DIAL. The identification of compounds involved comparing them to purified standards from a library and cross-referencing against several metabolomics databases, including METLIN, MassBank of North America (MoNA), Human Metabolome Database (HMDB), and LIPID MAPS. The MS/MS fragmentation of metabolites was then compared with candidate molecules from the databases and validated using earlier literature on similar compounds. The Metabolomics Center of Biocenter Kuopio also maintains an in-house library of over 1800 authenticated standards, which includes retention time, mass-to-charge ratio (m/z), and chromatographic data (including MS/MS spectral data) for all molecules present in the library.

### Sample Size Calculations and Statistical Analyses

We have described the sample size calculations for the PANIC study earlier [16]. Sample size calculations for the PANIC study were based on the effects of a dietary intervention on fasting serum insulin and Homeostatic Model Assessment for Insulin Resistance (HOMA-IR) among children in the Special Turku Coronary Risk Factor Intervention Project (STRIP) [15, 34]. Because of a larger number of children in the PANIC study than in the STRIP study, we approximated a slightly smaller difference for the change in fasting serum insulin and HOMA-IR of 0.3 SD between the intervention group (60% of children) and the control group (40% of children) with a power of 80% and a two-tailed p-value for the difference between the groups of 0.05, allowing for a 20% loss to follow-up or missing data. However, these calculations did not allow for non-independence within schools, and therefore the power could be lower than 80%. According to these calculations, we resulted in a sample size of at least 275 children in the intervention group and at least 183 children in the control group at baseline.

We analyzed the intervention effects on serum metabolites using the intention-to-treat principle and linear mixed-effects models adjusting for sex and age, including main effects for time and study group × time interaction, and assuming data missing at random. We considered results with a *p*-value of <0.05 statistically significant and also used false discovery rate corrected *p*-values (*q*-values) to control the results for a multiple testing error and help interpret the statistical significance of the results [35]. We used Cohen’s d values to assess effect sizes [36] and select serum molecular features for the identification of metabolites. We analyzed the data using the R software (R Core Team, 2021) and the IBM SPSS Statistics software (IBM Corp., Armonk, NY, USA).

## RESULTS

### Baseline Characteristics of Children

There were no differences in baseline characteristics between the intervention group and the control group, except the children in the intervention group consumed less high-fat milk and more low-fat milk than children in the control group **(Table 1)**.

**Table 1.** Baseline characteristics of children in the intervention group and the control group.

|  | Intervention<br>group<br>(N=296) | Control<br>group<br>(N=194) | *P*-value |
| --- | --- | --- | --- |
| Sex |  |  | 0.612 |
| Boys, $n$ (%) | 158 (53.4) | 99 (51.0) | |
| Girls, $n$ (%) | 138 (46.6) | 95 (49.0) | |
| Age, years | 7.6 (0.4) | 7.6 (0.4) | 0.329 |
| Body weight, kg | 26.8 (4.7) | 26.7 (5.3) | 0.788 |
| Body height, cm | 128.8 (5.5) | 128.5 (5.9) | 0.506 |
| Body height-SDS | 0.13 (0.99) | 0.11 (1.04) | 0.843 |
| BMI-SDS | -0.19 (1.04) | -0.22 (1.11) | 0.717 |
| Body weight status, $n$ (%) | | | 0.886 |
| Normal weight | 259 (87.5) | 172 (88.7) |  |
| Overweight | 27 (9.1) | 12 (6.2) |  |
| Obesity | 10 (3.4) | 10 (5.1) |  |
| Food consumption, g/day |  |  |  |
| Vegetables, fruit, and berries | 204 (115) | 221 (119) | 0.142 |
| High-fiber ($\geq 5\%$) grain products<sup>a</sup> | 63 (39) | 63 (40) | 0.862 |
| Low-fiber (<5%) grain products<sup>b</sup> | 113 (54) | 115 (51) | 0.620 |
| High-fat (60–80%) vegetable oil-based spreads | 6.9 (7.4) | 7.6 (8.5) | 0.393 |
| Vegetable oils | 4.3 (4.5) | 3.8 (3.9) | 0.283 |
| Butter and butter-based spreads | 5.7 (6.9) | 6.2 (7.2) | 0.447 |
| High-fat (≥1%) milk | 167 (208) | 220 (244) | 0.025 |
| Low-fat (<1%) milk | 411 (288) | 340 (288) | 0.014 |
| Red meat | 56 (29) | 58 (34) | 0.486 |
| Fish | 15 (20) | 16 (23) | 0.587 |
| Foods with high sugar content | 185 (136) | 206 (146) | 0.126 |
| Physical activity energy expenditure, kJ kg<sup>-1</sup> d<sup>-1</sup> | 101 (32) | 96 (34) | 0.076 |
| Light physical activity, h/d | 8.7 (1.8) | 8.4 (1.8) | 0.071 |
| Moderate physical activity, h/d | 1.6 (0.9) | 1.5 (0.9) | 0.162 |
| Vigorous physical activity, h/d | 0.4 (0.4) | 0.4 (0.4) | 0.716 |
| Sedentary time, excluding sleep, h/day | 3.7 (2.0) | 4.0 (2.3) | 0.362 |

The values are unadjusted means (standard deviations) and their *p* values from the independent samples T-test for continuous variables and numbers (percentages) and their *p* values from the Chi-square test for categorical variables.
Data on sex, age, pubertal status, body weight, body height, body height-SDS, BMI-SDS and body weight status were available for 306 children in the intervention group and for 198 children in the control group; on total physical activity energy expenditure for 282 children in the intervention group and for 184 children in the control group; on light, moderate and vigorous physical activity for 268 children in the intervention group and for 171 children in the control group; on sedentary time for 266 children in the intervention group and for 171 children in the control group; and on dietary factors for 246 children in the intervention group and for 163 children in the control group.
<sup>a</sup>Whole grain pasta, rice, and oatmeal
<sup>b</sup>White pasta, rice, and flour
<sup>c</sup>Sugar-sweetened beverages, fruit juice, candies, chocolate, added sugar, ice cream, puddings, pastries, and biscuits

### Effects of Diet and Physical Activity Intervention on Serum Metabolites

The intervention had statistically significant effects on 315 molecular features over the first two years (**Figure 2**), and among them, 80 metabolites were annotated (**Supplement 1**). Of these 80 metabolites, 17 were affected by the intervention over two and eight years, 59 only over two years, and three only over eight years (**Table 2**).

**Figure 2.**
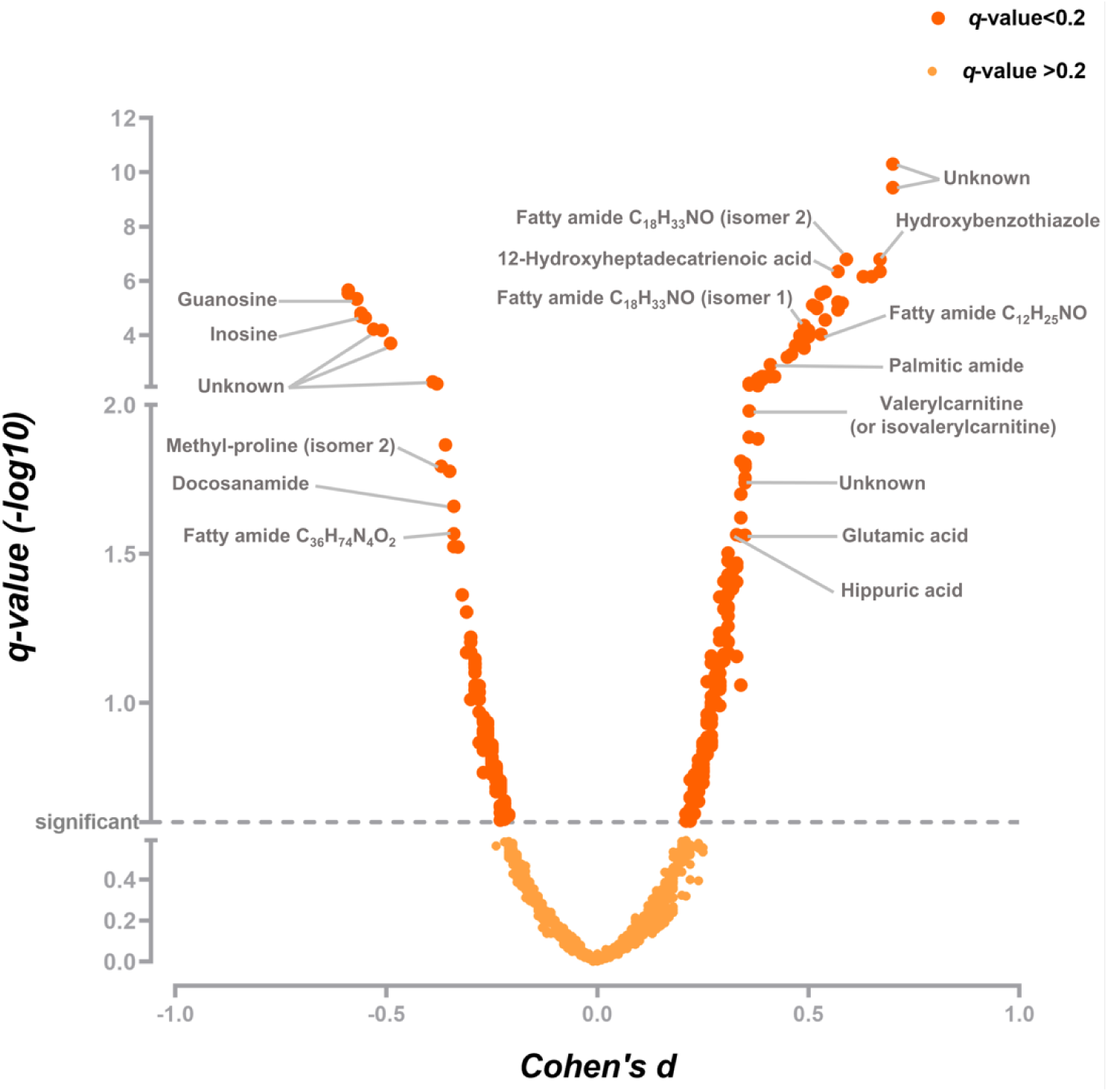
Effect sizes of the diet and physical activity intervention on 315 serum molecular features over two years adjusted for age and sex, a positive value indicating a positive effect of the intervention and a negative value indicating a negative effect of the intervention.

**Table 2.**
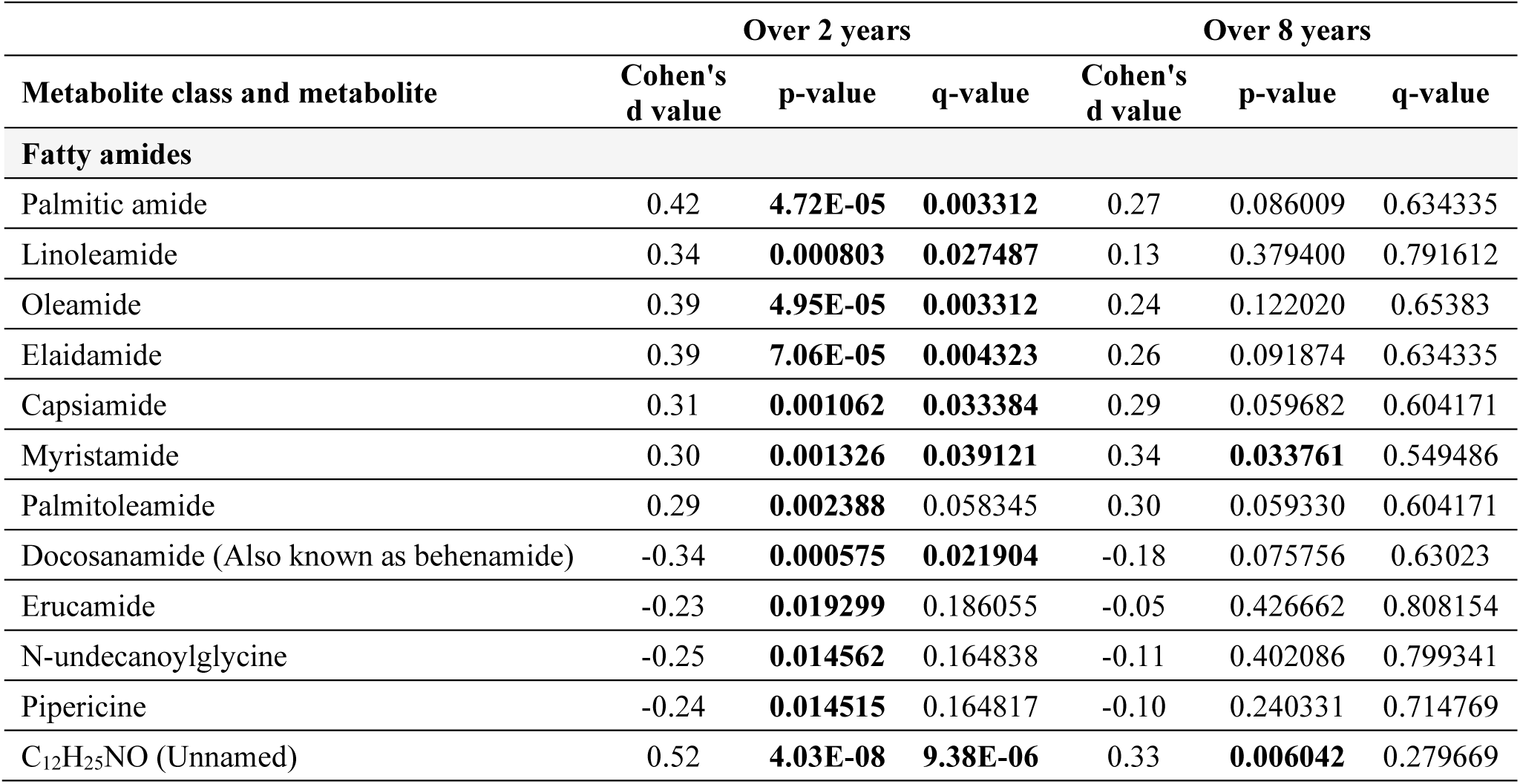
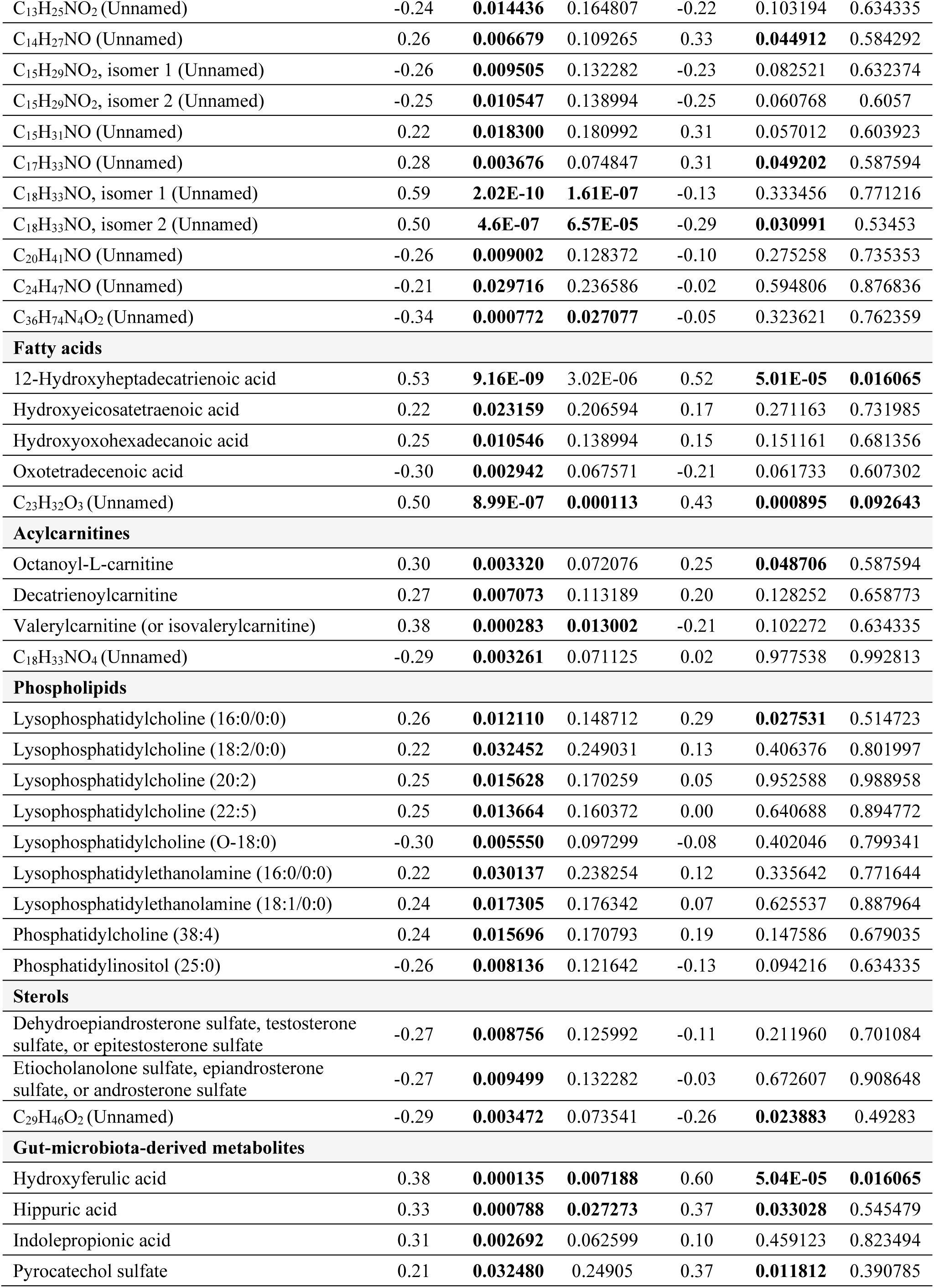
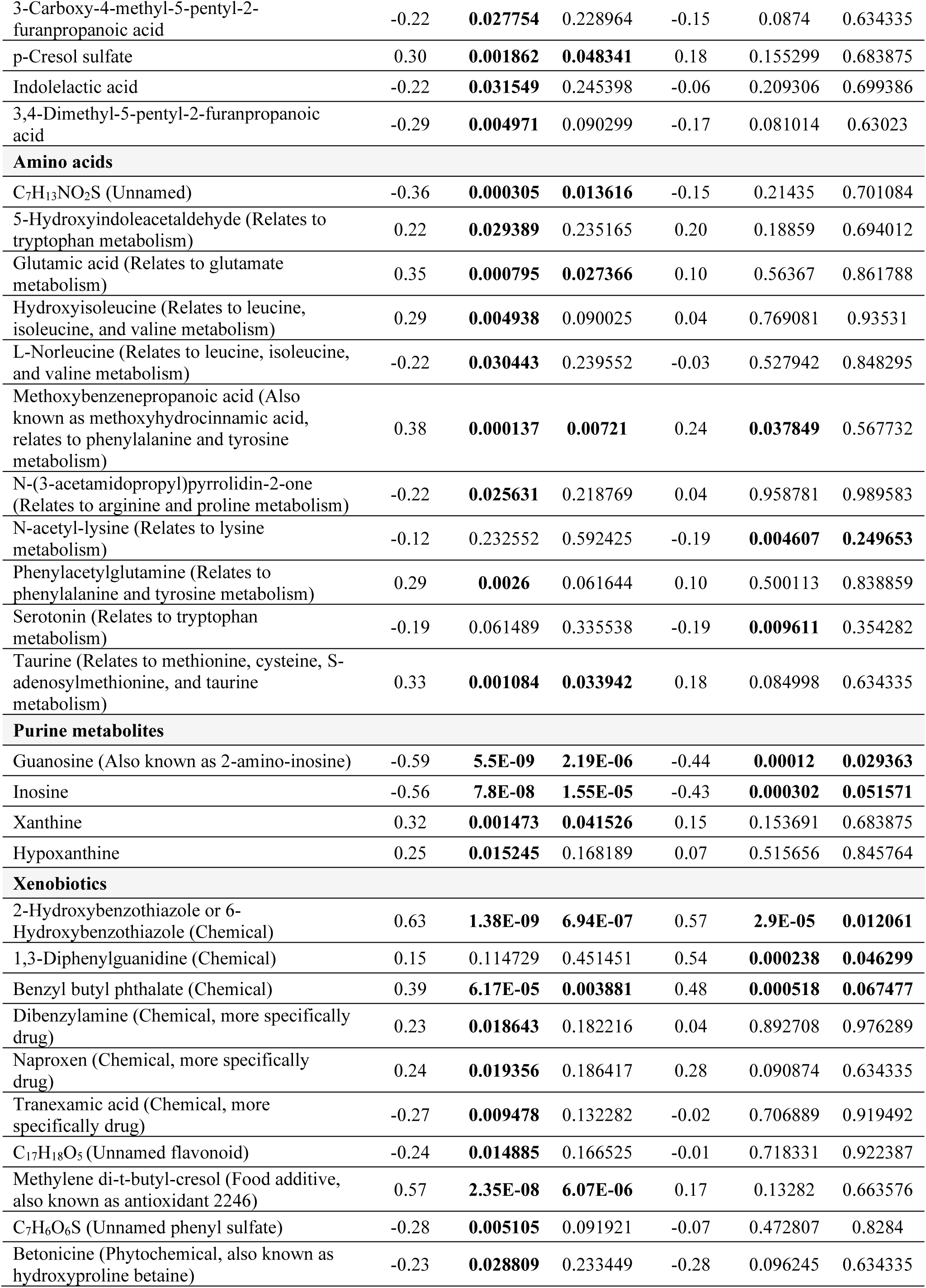
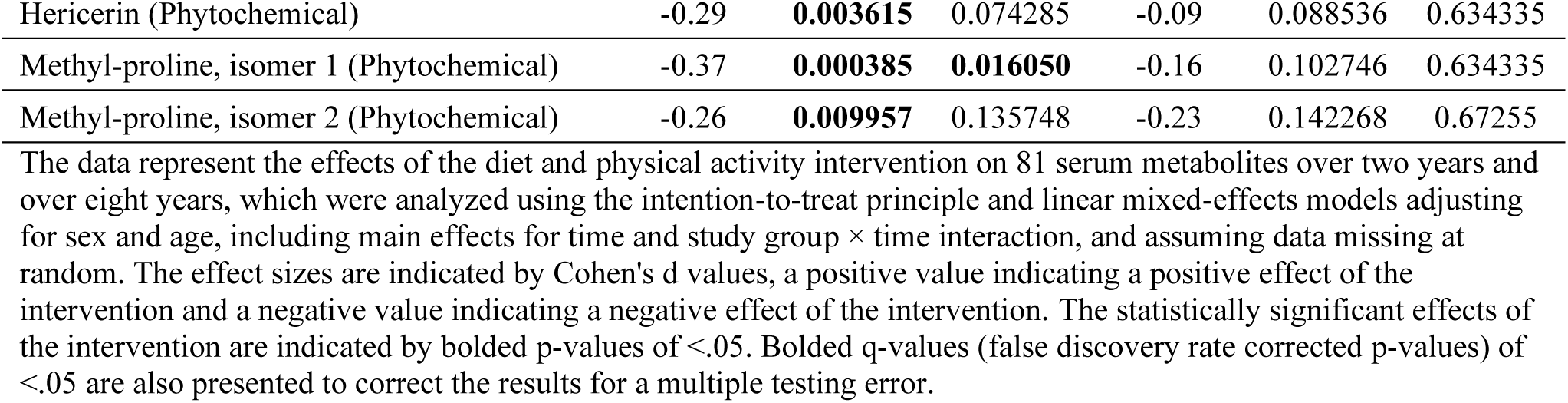
Effects of diet and physical activity intervention on serum metabolites over two and eight years.

The intervention had effects on 23 **fatty amides** (11 known, 12 unnamed) over two years (**Table 2**, **Figure 3**). Palmitic amide, linoleamide, oleamide, elaidamide, capsiamide, and myristamide decreased less in the intervention group than in the control group; palmitoleamide did not change in the intervention group but decreased in the control group; and docosanamide, erucamide, N-undecanoylglycine, and pipericine decreased in the intervention group but increased in the control group (**Figure 3**). The differences in all these known fatty amides, except N-undecanoylglycine, between the groups persisted until eight years (**Figure 3**), although the intervention effect on only myristamide remained statistically significant until then (**Table 2**).

**Figure 3.**
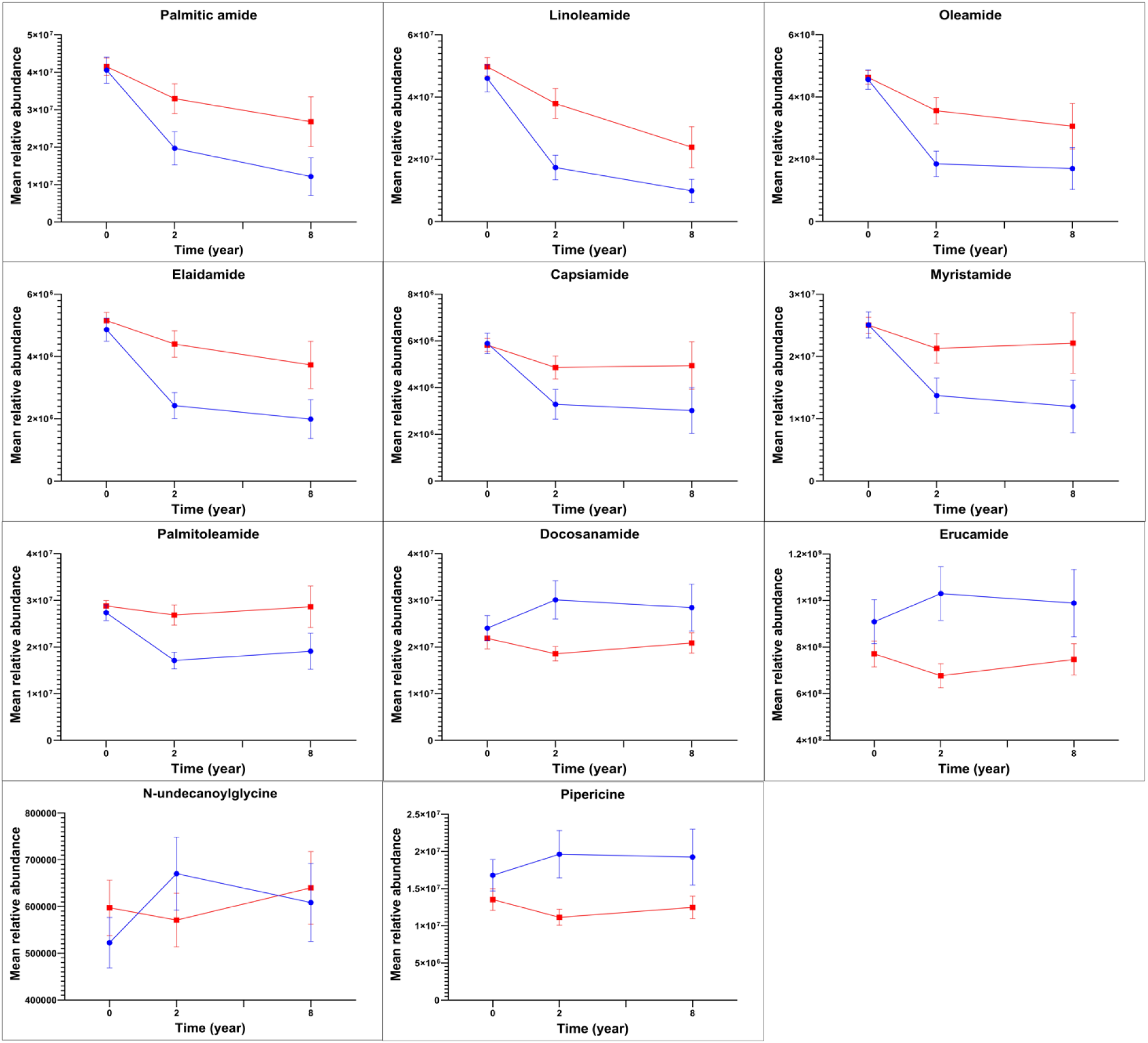
Mean relative abundances of serum fatty amides in the intervention group (red lines) and in the control group (blue lines) at baseline, 2-year follow-up, and 8-year follow-up examinations.

The intervention affected five unsaturated **fatty acids** (four known, one unnamed) over the first two years (**Table 2**, **Figure 4**). 12-hydroxyheptadecatrienoic acid and hydroxyeicosatetraenoic acid decreased less in the intervention group than in the control group; hydroxyoxohexadecanoic acid increased more in the intervention group than in the control group; and oxotetradecenoic acid decreased in the intervention group but increased in the control group (**Figure 4**). The differences in all these known fatty acids between the groups remained until eight years (**Figure 4**), although the intervention effect on only 12-hydroxyheptadecatrienoic acid persisted statistically significant until then (**Table 2**).

**Figure 4.**
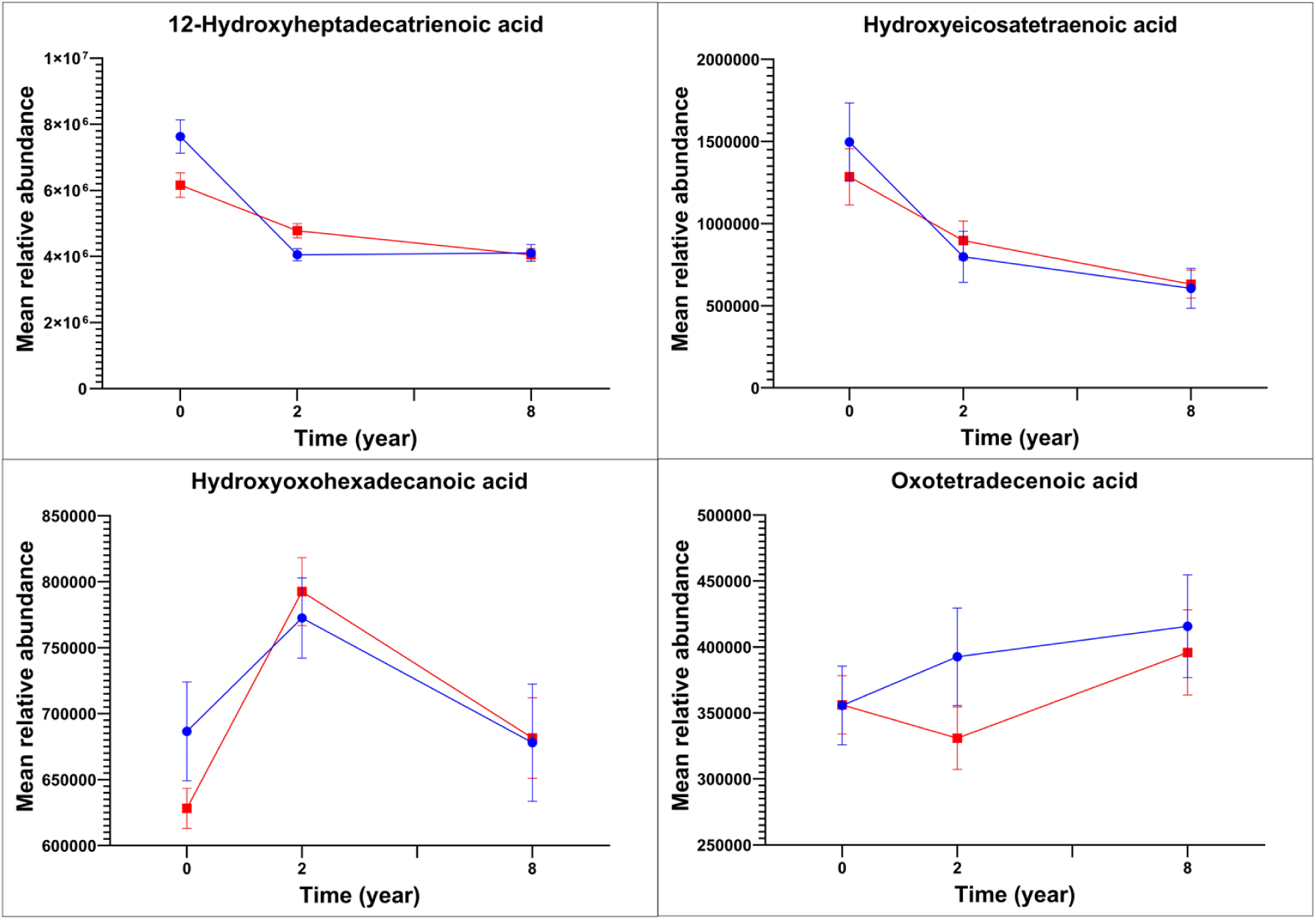
Mean relative abundances of serum fatty acids in the intervention group (red lines) and in the control group (blue lines) at baseline, 2-year follow-up, and 8-year follow-up examinations.

The intervention had effects on four **acylcarnitines** (three known, one unnamed) over the first two years (**Table 2**, **Figure 5**). Octanoyl-L-carnitine, decatrienoylcarnitine, and valerylcarnitine (or isovalerylcarnitine) increased in the intervention group and decreased in the control group (**Figure 5**). The differences in octanoyl-L-carnitine and decatrienoylcarnitine between the groups persisted until eight years (**Figure 5**), although the intervention effect on only octanoyl-L-carnitine remained statistically significant until then (**Table 2**).

**Figure 5.**
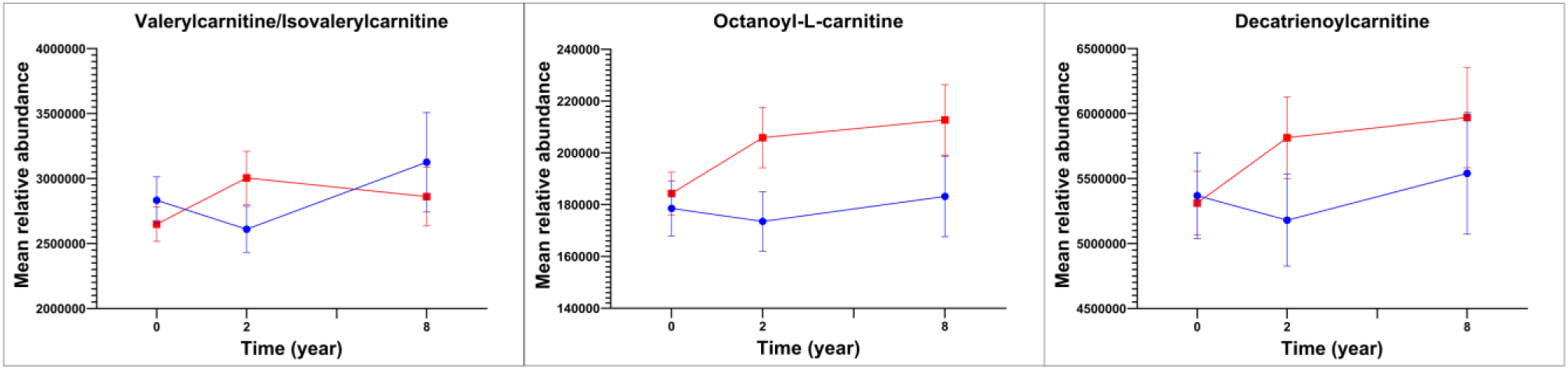
Mean relative abundances of serum acylcarnitines in the intervention group (red lines) and in the control group (blue lines) at baseline, 2-year follow-up, and 8-year follow-up examinations.

The intervention affected nine **phospholipids** (all nine known) over the first two years (**Table 2**, **Figure 6**). Lysophosphatidylcholine(16:0), lysophosphatidylcholine(18:2), lysophosphatidylcholine(20:2), lysophosphatidylcholine(22:5), phosphatidylcholine(38:4), and lysophosphatidylethanolamine(18:1) increased more in the intervention group than in the control group; lysophosphatidylethanolamine(16:0) decreased less in the intervention group than in the control group; and lysophosphatidylcholine(O-18:0) and phosphatidylinositol(25:0) decreased more in the intervention group than in the control group; (**Figure 6**). The differences in lysophosphatidylcholine(16:0), lysophosphatidylcholine(18:2), phosphatidylcholine(38:4), lysophosphatidylcholine(O-18:0), and phosphatidylinositol(25:0) between the groups remained until eight years (**Figure 6**), although the intervention effect on only lysophosphatidylcholine(16:0) persisted statistically significant until then (**Table 2**).

**Figure 6.**
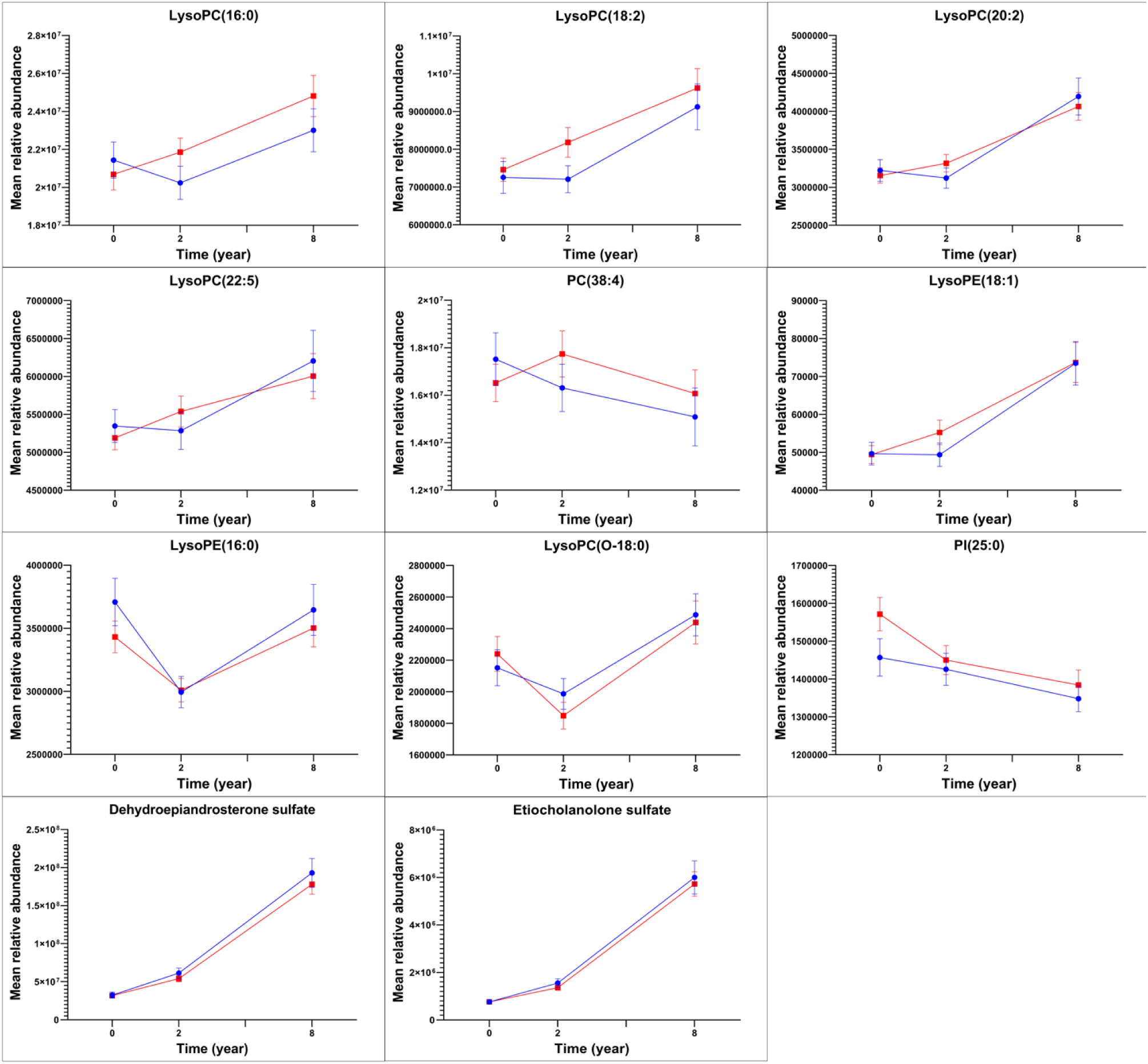
Mean relative abundances of serum phospholipids and sterols in the intervention group (red lines) and in the control group (blue lines) at baseline, 2-year follow-up, and 8-year follow-up examinations.

The intervention had effects on three **sterols** (two known, one unnamed) over the first two years (**Table 2**, **Figure 6**). Dehydroepiandrosterone sulfate (or epitestosterone sulfate or testosterone sulfate) and etiocholanolone sulfate (or epiandrosterone sulfate or androsterone sulfate) increased slightly less in the intervention group than in the control group (**Figure 6**). The differences in dehydroepiandrosterone sulfate and etiocholanolone sulfate between the groups persisted until eight years (**Figure 6**), although the intervention effects on these sterols did not remain statistically significant until then (**Table 2**).

The intervention had effects on eight **gut-microbiota-derived metabolites** (all eight known) over the first two years (**Table 2**, **Figure 7**). Hydroxyferulic acid, hippuric acid, indolepropionic acid, and pyrocatechol sulfate did not change in the intervention group but decreased in the control group; 3-carboxy-4-methyl-5-pentyl-2-furanpropanoic acid did not change in the intervention group but increased in the control group; *p*-cresol sulfate increased in the intervention group but decreased in the control group; indolelactic acid increased less in the intervention group than in the control group; and 3,4-dimethyl-5-pentyl-2-furanpropanoic acid decreased in the intervention group but increased in the control group (**Figure 7**). The differences in hydroxyferulic acid, hippuric acid, pyrocatechol sulfate, 3-carboxy-4-methyl-5-pentyl-2-furanpropanoic acid, indolelactic acid, and 3,4-dimethyl-5-pentyl-2-furanpropanoic acid between the groups remained until eight years (**Figure 7**), although the intervention effects on only hydroxyferulic acid, hippuric acid, and pyrocatechol sulfate persisted statistically significant until then (**Table 2**).

**Figure 7.**
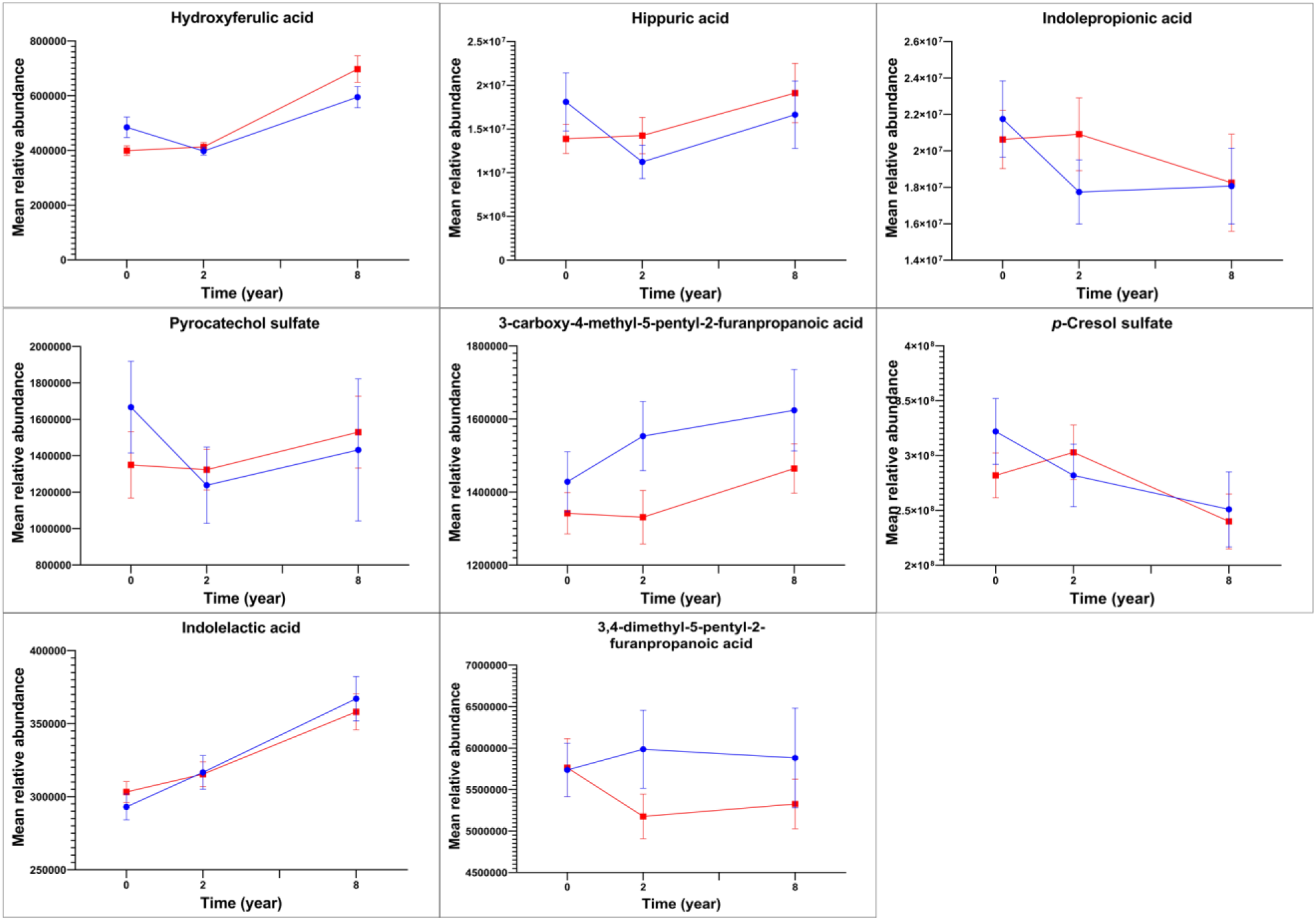
Mean relative abundances of serum gut-microbiota-derived metabolites in the intervention group (red lines) and in the control group (blue lines) at baseline, 2-year follow-up, and 8-year follow-up examinations.

The intervention had effects on 11 **amino acids** (10 known, one unnamed) over the first two years (**Table 2**, **Figure 8**). Methoxybenzenepropanoic acid increased more in the intervention group than in the control group; glutamic acid did not change in the intervention group but decreased in the control group; taurine decreased less in the intervention group than in the control group; and hydroxyisoleucine increased in the intervention group but decreased in the control group (**Figure 8**). The difference in methoxybenzenepropanoic acid between the groups persisted until eight years (**Figure 8**), and the intervention effect on this amino acid remained statistically significant until then (**Table 2**).

**Figure 8.**
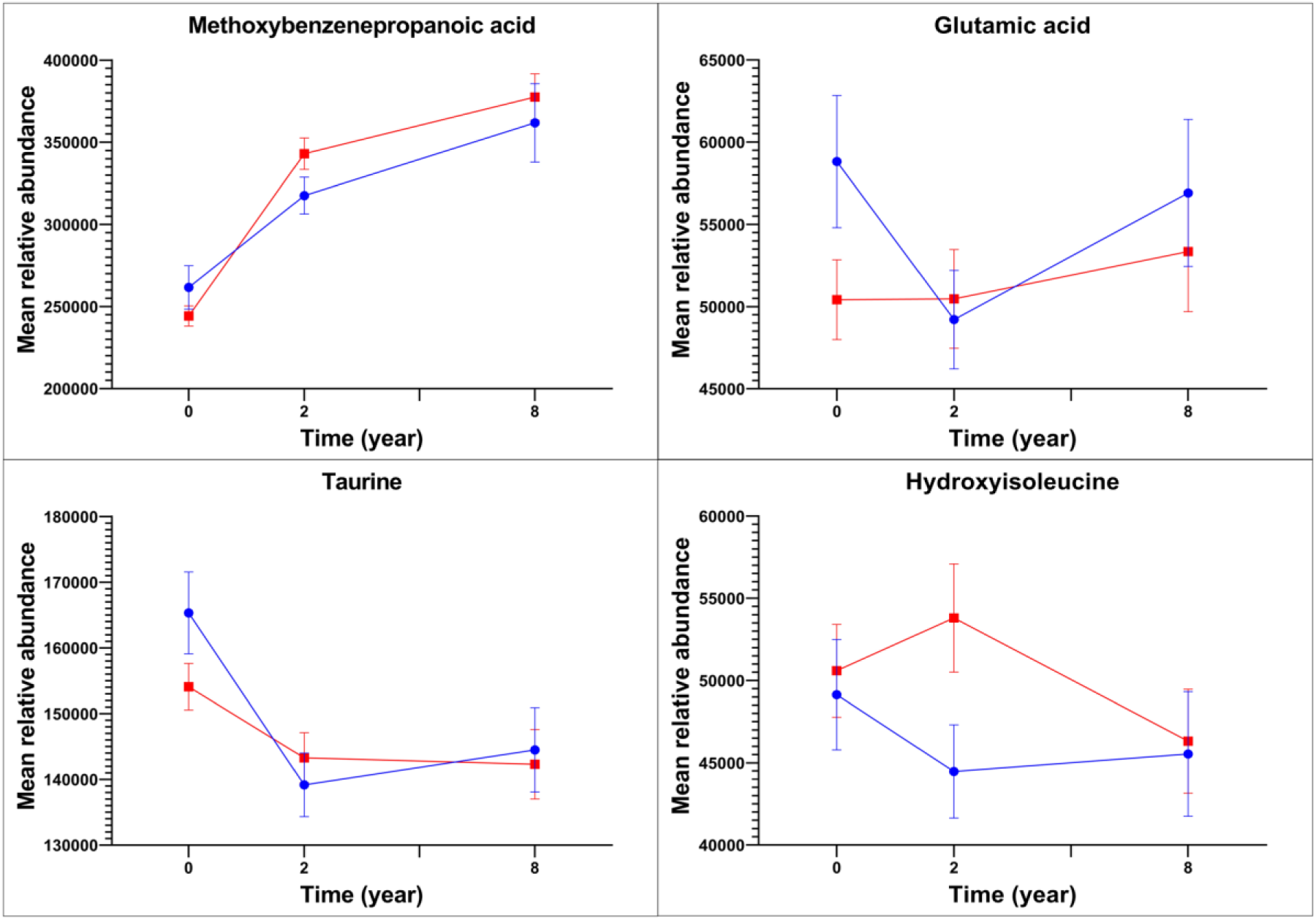
Mean relative abundances of serum amino acids in the intervention group (red lines) and in the control group (blue lines) at baseline, 2-year follow-up, and 8-year follow-up examinations.

The intervention affected four **purine metabolites** (all known) over the first two years (**Table 2**, **Figure 9**). Guanosine and inosine decreased in the intervention group but increased in the control group, whereas xanthine and hypoxanthine decreased less in the intervention group than in the control group (**Figure 9**). The differences in guanosine and inosine between the groups persisted until eight years (**Figure 9**), and the intervention effects on these purine metabolites remained statistically significant until then (**Table 2**).

**Figure 9.**
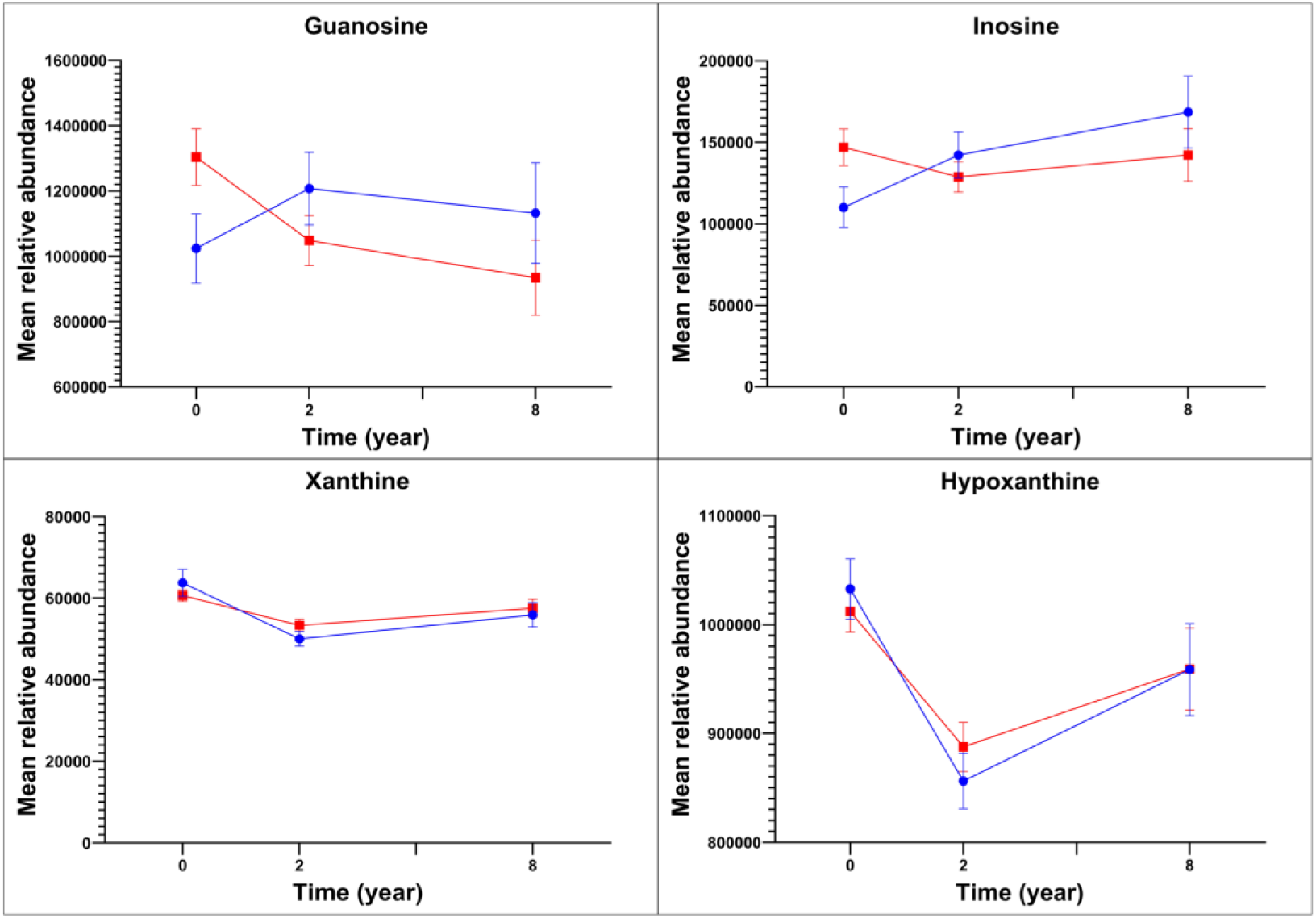
Mean relative abundances of serum purine metabolites in the intervention group (red lines) and in the control group (blue lines) at baseline, 2-year follow-up, and 8-year follow-up examinations.

## DISCUSSION

This 8-year lifestyle intervention study in a general population of children showed that the combined diet and PA intervention had effects on numerous serum metabolites that could influence cardiometabolic and overall health since childhood. The effects of the diet and PA intervention were most pronounced on serum fatty amides, but the intervention also affected other potentially important serum lipids, including fatty acids, acylcarnitines, phospholipids, and sterols, as well as serum gut-microbiota-derived metabolites, amino acids, and purine metabolites.

Our diet and PA intervention had effects on several serum fatty amides, which are bioactive lipid signaling molecules with a variety of cardiometabolic and other physiological functions, including sleep induction, pain and anxiety control, eating behavior, weight control, angiogenesis, arterial dilation, as well as anti-inflammatory, neuroprotective, and anticonvulsive effects, and whose functions depend on the length and degree of unsaturation of their acyl group [37, 38]. The diet and PA intervention attenuated the decrease of serum levels of several short-chain primary fatty amides with 16-18 carbons, including palmitic amide, linoleamide, oleamide, elaidamide, capsiamide, and myristamide, all of which are produced endogenously [37, 39]. This finding suggests that the diet and PA intervention may have directly affected fatty amide metabolism, which could have been be through its effects on fatty acid amide hydrolase activity [40]. This enzyme is known to modulate circulating fatty amide concentrations and regulate several physiological processes in the human body [41, 42]. However, many of these short-chain fatty amides, such as palmitic amide, linoleamide, and oleamide, are not only produced endogenously but are also rich in vegetable oils [38], suggesting that the effects of our diet and PA intervention on the serum levels of these fatty amides could be partly explained by the increased consumption of vegetable oils in the intervention group [17, 24]. The current result with regard to the effect of our diet and PA intervention on serum linoleamide concentration is consistent with our earlier finding that the same lifestyle intervention increased plasma levels of linoleic acid [18], which is metabolically related to linoleamide.

Moreover, a single bout of exercise has been shown to increase oleamide concentrations in the skeletal muscle of rats [43], and on the other hand, gut-microbiota-produced fatty amides have been shown to mediate the gut-brain feedback loop related to motivation for exercise in mice [44]. In contrast to the effects of our diet and PA intervention on serum short-chain primary fatty amides, serum levels of two long-chain primary fatty amides with 20-22 carbons, docosanamide and erucamide, decreased in the intervention group but increased in the control group. These long-chain fatty amides have not been found to be synthesized in the human body but have been shown to be exogenous compounds [45]. However, docosanoic acid and erucic acid, which are fatty acids metabolically related to docosanamide and erucamide, are abundant in rapeseed oil, canola oil, and olive oil, which are common vegetable oils in Finland. Thus, one explanation for the effects of our diet and PA intervention on serum levels of docosanamide and erucamide may be that these fatty amides are gut-microbiota-derived metabolites [46] of dietary fats [47].

We observed that the diet and PA intervention affected serum levels of unsaturated fatty acids for example by attenuating the decrease of hydroxyheptadecatrienoic acid and hydroxyeicosatetraenoic acid, increasing hydroxyoxohexadecanoic acid, and decreasing oxotetradecenoic acid. These results agree with our previous finding that the diet and PA intervention increased the consumption of vegetable oil–based margarine [42] and plasma level of linoleic acid [17], which is a precursor of hydroxyheptadecatrienoic acid, and increased plasma levels of α-linolenic acid and docosapentaenoic acid in the present cohort of children [18].

We found that the diet and PA intervention increased serum levels of a few short-chain and medium-chain acylcarnitines, including valerylcarnitine (or isovalerylcarnitine), octanoylcarnitine, and decatrienoylcarnitine. Acylcarnitines transport fatty acids from the cytoplasm into the mitochondria to produce energy in beta-oxidation [48, 49]. During fasting or exercise, the body may increase the breakdown of fatty acids and the production of acylcarnitines to provide additional energy for skeletal muscles [50]. PA has been reported to spike up circulating short- and medium-chain acylcarnitines and thereby trigger fatty acid oxidation in adults [51]. Increased fructose intake as a result of increased fruit or fruit juice consumption has also been found to increase short-chain acylcarnitines and decrease medium- and long-chain acylcarnitines in circulation [52, 53]. We have previously shown that the lifestyle intervention increased PA and fruit consumption in the present cohort of children [24] that may partly explain our present finding that the lifestyle intervention increased serum levels of short-chain acylcarnitines among children.

In addition to the effects of our diet and PA intervention on serum levels of several fatty amides, acylcarnitines, and fatty acids, the lifestyle intervention also affected serum levels of many phospholipids and sterols, which provides evidence for the broad impacts of our lifestyle intervention on lipid metabolism in a general population of children. These findings on lipid metabolism are also consistent with our earlier observations dealing with the beneficial effects of our diet and PA intervention on diet quality [23, 42], plasma fatty acids [18], and plasma LDL cholesterol [17] among children. Moreover, the present observation that serum dehydroepiandrosterone sulfate, measured by non-targeted LC-MS metabolic profiling, increased less in the intervention group than in the control group agrees with our recent finding, using a targeted LC-MS analysis, that the same diet and PA intervention attenuated the increase of serum dehydroepiandrosterone and dehydroepiandrosterone sulfate in both sexes and also attenuated the increase of serum androstenedione and testosterone and delayed pubarche in boys [54].

Our diet and PA intervention had effects on serum levels of several gut-microbiota-derived metabolites, which are chemically phenolic acids, amino acids, or fatty acids. The intervention attenuated the decrease of hydroxyferulic acid, hippuric acid, indolepropionic acid, and pyrocatechol sulfate, increased *p*-cresol sulfate, and attenuated the increase of indolelactic acid. Urinary hippuric acid has been proposed a biomarker for fruit and vegetable consumption in children [55] and for polyphenol intake in adults [56]. Higher serum level of hippuric acid has been associated with a lower fasting blood glucose concentration and higher insulin secretion among adults [20]. A higher dietary intake of fiber has been associated with a higher serum level of indolepropionic acid [57] that has been related to higher insulin secretion and a lower risk of type 2 diabetes among adults with impaired glucose tolerance [57, 58]. We have earlier found that the diet and PA intervention increased vegetable consumption and fiber intake [23, 24] and attenuated the increase of insulin resistance [16] in the present cohort of children. All these observations together suggest that gut-microbiota-derived metabolites, such as hippuric acid and indolepropionic acid, could mediate the beneficial effects of improved diet quality on glucose metabolism among adults and children. Moreover, our intervention decreased serum levels of two furan fatty acids, 3,4-dimethyl-5-pentyl-2-furanpropanoic acid and 3-carboxy-4-methyl-5-pentyl-2-furanpropanoic acid, which have not been reported to be engogenous compounds but have been found in fish and regarded as biomarkers for fish consumption [59–61]. However, as we have previously observed no difference in fish consumption between the intervention group and the control group [24], eating fish is unlikely to explain the intervention effects on these furan fatty acids. Instead, furan fatty acids have been suggested to be derived from gut microbiota [59], and the effects of our lifestyle intervention on these furan fatty acids could be partly due to changes in gut microbiota.

We found that the diet and PA intervention had effects on serum levels of several amino acids for example by attenuating the decrease of taurine, increasing hydroxyisoleucine, and preventing the decrease of glutamic acid. Taurine has been shown to slow aging process and increase in response to PA [62]. Four-hydroxyisoleucine has been found to inhibit obesity-related insulin resistance in adipocytes and hepatocytes by reducing inflammation and regulating the state of M1/M2 macrophages [63]. Glutamic acid is metabolized in the human body into glutamate, which is among key neurotransmitters in the central nervous system and has been suggested to be important for learning and memory [64]. Therefore, these amino acids may be molecular mechanisms underlying the beneficial effects of our lifestyle intervention on cardiometabolic and brain health among children.

Our diet and PA intervention affected purine metabolism by decreasing serum levels of guanosine and inosine and by increasing serum levels of hypoxanthine and xanthine. Purine metabolism is essential for adequate adenosine triphosphate resynthesis from degraded bases and the preservation of skeletal muscle adenine nucleotide pool for energy metabolism during strenuous exercise [65].

High-intensity speed-power exercise training has been shown to activate the purine salvage pathway [66], in which hypoxanthine and guanine are converted to inosine monophosphate and guanosine monophosphate for adenine nucleotide resynthesis [65]. Our findings suggest that the diet and PA intervention, which did not include high-intensity speed-power exercise, did not activate the purine salvage pathway as hypoxanthine increased and guanosine and inosine decreased in response to the intervention. In the purine degradation pathway, accumulated adenosine monophosphate is degraded to inosine and hypoxanthine, the latter of which is further metabolized to xanthine and uric acid by xanthine dehydrogenase and xanthine oxidase [67]. However, circulating hypoxanthine levels are more affected by the inhibition of the purine salvage pathway and the activation of liver xanthine oxidase than by the activation of the purine degradation pathway [68]. It would have been useful to be able to measure xanthine oxidase activity from skeletal muscle, the liver, or adipose tissue to interpret the effects of our diet and PA intervention on purine metabolism and cardiometabolic health as increased xanthine oxidase activity has been associated with obesity, liver dysfunction, hyperuricemia, dyslipidemia, and insulin resistance by increasing reactive oxygen species [69]. Finally, gut microbiota has been found to affect purine metabolites, especially guanosine, inosine, and hypoxanthine [70], that could aprtly explain the effects of our lifestyle intervention on purine metabolites.

We had for the first time an opportunity to study the long-term effects of a diet and PA intervention on a wide spectrum of serum metabolites using the nontargeted metabolomics analysis by the sensitive LC-MS method in a general population of children followed up until adolescence. The advantage of studying children instead of adults is that it decreases confounding by chronic diseases, medications, smoking, and alcohol consumption. A limitation of our study is that we did not randomly allocate the children into the intervention group and the control group. However, we analyzed the data using linear mixed-effects models that allowed us to control for confounding by possible differences in serum metabolites at baseline. Puberty is a potential confounding factors in studies among children, as it markedly affects metabolism. However, puberty is unlikely to be a confounding factor in our study as we included only prepubertal children at baseline in the statistical analyses and there was no difference in the occurrence of puberty between the intervention group and the control group during the study [16]. Many effects of the diet and PA intervention on serum metabolites found over the first two years were no longer statistically significant after eight years, although small differences in these metabolites between the groups still remained. This is likely to be explained by a modest intensity of the lifestyle intervention after the first two years and a limited statistical power in the statistical analyses due to a reduced number of participants attending the 8-year follow-up examinations. Finally, it would have been useful to have data on fecal microbial composition to provide a more comprehensive insight into the effects of the diet and PA intervention on gut-microbiota-derived metabolites during childhood and adolescence.

## Conclusions

This long-term diet and PA intervention study demonstrates that the nontargeted metabolomics analysis of serum samples using the LC-MS method increases our understanding of possible mechanisms underlying the beneficial effects of lifestyle interventions on cardiometabolic and overall health during childhood and adolescence and thereby provides novel and valuable insights into the prevention of cardiometabolic and other non-communicable diseases since childhood. Other long-term diet and PA intervention studies using the LC-MS metabolomics and other omics analyses are warranted to provide further evidence on these mechanisms that can be used in the early prevention of non-communicable diseases.

## Data Availability

All data produced in the present study are available upon reasonable request to the authors

https://submit.medrxiv.org/submission/download?msid=MEDRXIV/2024/305105&roleName=author&gotoPageName=proof&continueToPage=proof&supp=true

## List of abbreviations

PA: physical activity
LC-MS: liquid chromatography - mass spectrometry
PANIC: Physical Activity and Nutrition in Children

## Declarations

### Ethics approval and consent to participate

The Research Ethics Committee of the Hospital District of Northern Savo approved the study protocol in 2006 and 2015 (Statements 69/2006 and 422/2015). The caregivers gave their written informed consent, and the children provided their assent to participation. Moreover, the caregivers and adolescents gave their written informed consent before 8-year examinations.

### Competing interests

KH, VMK, and AK are affiliated with Afekta Technologies Ltd <u>Afekta Technologies Ltd. - Metabolomics service company</u>. Other authors of this article declare no competing interests.

### Funding

The PANIC study has been supported financially by grants from the Academy of Finland, the Ministry of Education and Culture of Finland, the Ministry of Social Affairs and Health of Finland, the Research Committee of the Kuopio University Hospital Catchment Area (State Research Funding), Finnish Innovation Fund Sitra, Social Insurance Institution of Finland, Finnish Cultural Foundation, Foundation for Pediatric Research, Diabetes Research Foundation in Finland, Finnish Foundation for Cardiovascular Research, Juho Vainio Foundation, Paavo Nurmi Foundation, Yrjö Jahnsson Foundation, and the city of Kuopio. Moreover, the current work was supported by Horizon 2020 research and innovation programme of European Union (Grant 874739 for LongITools project) to MK, TAL, and KH. The funding bodies have no role in the design of the study and collection, analysis, and interpretation of data and in writing the manuscript.

### Authors’ contributions

TAL as the principal investigator of the PANIC study has access to all of its data and takes responsibility for the integrity of the data and the accuracy of the data analyses. IZ, AME, AK, JV, ML, TS, EAH, NL, SS, US, SA, KH, and TAL contributed to the acquisition of the data for the current work. TAL, KH, and MK secured funding for the work. TAL and KH conceptualized and designed the current work and contributed equally to it as senior authors. AK and SM performed the statistical analyses of the work. IZ, TAL, and KH drafted the manuscript, and all other authors critically revised the manuscript for its intellectual content. TAL, KH, MK, ML, and SA provided their administrative, technical, or material support for the work.

## Acknowledgements

We are indebted to all members of the PANIC research team for their invaluable contribution to data acquisition and to all families participating in the PANIC study. The authors would also like to thank Biocenter Finland (www.biocenter.fi) and Biocenter Kuopio (www.uef.fi/web/bck) for supporting the study.

## Notes

### Competing Interest Statement

KH, VMK, and AK are affiliated with Afekta Technologies Ltd Afekta Technologies Ltd. Metabolomics service company. Other authors of this article declare no competing interests.

### Clinical Trial

ClinicalTrials.gov NCT01803776

### Clinical Protocols

https://www.panicstudy.fi/

### Author Declarations

Ethics approval and consent to participate: The Research Ethics Committee of the Hospital District of Northern Savo approved the study protocol in 2006 and 2015 (Statements 69/2006 and 422/2015). The caregivers gave their written informed consent, and the children provided their assent to participation. Moreover, the caregivers and adolescents gave their written informed consent before 8-year examinations.

